# EasyCog: a fast and effective numerical tool for the self-assessment of cognitive functions

**DOI:** 10.64898/2026.09.23.26363767

**Authors:** Sarah Lejczak, Lisa Hadri, Xavier Fabian, Ophélie Martinie, Grégory Justin, Ségolène Lithfous, Eric Salvat, Julien Nizard, Pierrick Poisbeau

**Author notes:** Corresponding author:* Prof. Pierrick Poisbeau, Cognitive and Adaptive Neuroscience Laboratory (CNRS UMR 7364), 12 rue Goethe, 67000 Strasbourg, France. Data are available upon request. These authors contributed equally (Sarah Lejczak, Lisa Hadri).

## Abstract

Many pathological conditions, including chronic pain (CP), can impair cognitive functions. These functions are essential for adapting and evolving in concordance with the environment, which are key processes for coping with CP. However, in CP centers, cognitive assessment is rarely included in CP patient follow-up due to a lack of time, appropriate tools, or trained health professionals. To fill this gap, our team developed EasyCog, a digital tool that enables rapid self-assessment of cognitive functions. It enables cognitive evaluation without an experimenter’s help, thanks to speech recognition powered by an artificial neural network. In the initial validation steps, we established normative values for 7 cognitive functions on a cohort of 76 healthy subjects. Because EasyCog includes tests comparable to the Montreal Cognitive Assessment (MoCA) and the Mini-Mental State Examination (MMSE), we assessed its reliability against available normative values. To assess its specificity, we compared data from healthy subjects with 15 subjects with mild cognitive impairment (MCI) and 15 age-matched CP patients selected from a medical center. Finally, we tested EasyCog’s test-retest reliability by repeating sessions at different time intervals. EasyCog showed excellent sensitivity to age-related decline, similar to MoCA normative values. Several cognitive functions were significantly and differentially impaired in MCI subjects and CP patients compared to normative values in healthy subjects, confirming good specificity. Finally, test-retest analysis showed no effect at a 4-month interval. In conclusion, EasyCog appears to be a convenient and reliable numeric tool for rapidly assessing and monitoring cognitive functions in patients.

## 1. Introduction

Cognitive functions enable individuals to adapt to and effectively engage with their environment. These functions naturally decline across the lifespan (Jost & Kujach, 2025; Klimova, 2025), a process that is accelerated under certain pathological conditions. For instance, cognitive decline is particularly pronounced in neurodegenerative diseases (e.g., Alzheimer’s disease, amyotrophic lateral sclerosis) and acquired brain injury, and is associated with reduced autonomy and poorer quality of life (Tait et al., 2019; Tsermentseli et al., 2012). While these neurological conditions lead to severe cognitive deficits, other disorders may affect cognition more subtly. Psychiatric disorders (e.g., schizophrenia, depression) and chronic pain (CP) have been associated with cognitive impairments that meet criteria for mild cognitive impairment (MCI) (Corti et al., 2021; Moriarty et al., 2011). MCI is commonly defined as “an acquired, significant, and progressive decline in one or more cognitive domains, with preserved independence in activities of daily living” (French recommendations by Haute Autorité de Santé). Early identification of subtle cognitive impairments is therefore important for patients’ quality of life.

In chronic conditions such as CP (i.e., persistent or recurrent pain lasting more than 3 months), even subtle cognitive impairment represents an additional clinical burden. CP has been associated with impairments in several cognitive domains, including executive functions, attention, and memory (Berginström et al., 2024; Liu et al., 2014; Moriarty et al., 2011; Yoshino et al., 2021). In addition, pharmacological treatments commonly prescribed for CP, such as opioids and antiepileptics, may further contribute to cognitive dysfunction (Salinsky et al., 2010; Sjøgren et al., 2005). Cognitive functions are critical for the daily management of pain and for maintaining quality of life. They play a key role in treatment engagement and adherence, particularly for non-pharmacological and multidisciplinary interventions commonly used in pain management settings (Berginström et al., 2024). For instance, cognitive-behavioral therapy requires self-regulatory resources, closely linked to executive functioning, to maintain engagement and support the implementation and maintenance of adaptive behavioral strategies in daily life (Letzen et al., 2019; Solberg Nes et al., 2009). However, cognitive deficits are rarely identified in routine clinical practice, partly due to the lack of assessment tools and training among healthcare professionals in CP centers.

CP, which affects approximately 42% of the adult population in France (Fondation-Analgesia, 2025), represents a complex condition requiring a biopsychosocial approach, in which cognitive assessment is of relevance. In general clinical settings, where neuropsychologists are not available, brief cognitive screening tools such as the Montreal Cognitive Assessment [MoCA; (Nasreddine et al., 2005)] and the Mini-Mental State Examination [MMSE; (Folstein et al., 1975)] are commonly used by medical doctors. However, these instruments may present certain practical and methodological limitations in specific contexts. Although widely used and well validated, they are relatively time-consuming in routine practice, require clinician administration, and may be less suitable for repeated assessments over short intervals (Aiello et al., 2024; Gross et al., 2018). Furthermore, their reliance on cut-off scores remains debated, particularly for identifying mild cognitive impairment (MCI) (Carson et al., 2018). As the Montreal Cognitive Assessment (MoCA) and the Mini-Mental State Examination (MMSE) were originally developed for dementia screening, their thresholds may be less sensitive to the more subtle cognitive changes observed in conditions such as CP.

The aim of the present study was to validate a rapid, self-administered cognitive screening tool suitable for routine clinical use while maintaining robust psychometric properties. Unlike traditional clinician-administered instruments, this tool is delivered via a digital tablet and incorporates artificial intelligence (AI)- powered voice recognition to analyze the user’s verbal responses, thereby enabling self-administration and reducing examiner-related variability. This article describes the validation of the tool, including its sensitivity, test–retest reliability, and specificity. Normative data were obtained from a sample of healthy subjects (HS) aged from 18 to 93 years. In addition, specificity analyses were conducted by comparing performance across age groups in individuals with MCI or CP.

## 2. Materials and methods

### 2.1. Participants

The validation study of EasyCog began in January 2023, and 111 volunteers were recruited. Participants were contacted through advertisements at the University of Strasbourg, via websites and social media.

Sampling aimed to achieve a balanced distribution of sex and an adult age distribution centered around 55 years, based on previous validation studies of the Montreal Cognitive Assessment (MoCA) and the Mini-Mental State Examination (MMSE).

Undiagnosed volunteers were subsequently assigned to either the control group (healthy subjects, n = 76) if their MoCA score was ≥ 26/30, or to the MCI group (n = 15) if their score was < 26.

For the control group, non-inclusion criteria included: history of CP (i.e. persistent or recurrent pain lasting more than 3 months); neurological disorders or sequelae; psychiatric disorders; history of head injury with loss of consciousness or stroke; use of medications likely to affect cognition (e.g. antidepressants, antiepileptics, hypnotics), chronic use of analgesics (e.g. opioids, antidepressants, antiepileptics); use of psychoactive substances (e.g. amphetamines, opiates, cannabis, cocaine, benzodiazepines, barbiturates); belonging to a protected population (e.g. minors, pregnant women, individuals under legal guardianship); inability to provide informed consent; severe uncorrected hearing or visual impairments.

Participants with CP (CP patients; n = 15) were recruited through referrals from general practitioners specializing in pain management or during consultations in chronic pain centers. Eligible patients were adults aged 18 years or older with a diagnosis of chronic pain, whether newly referred or already receiving follow-up care, and whether or not they were receiving pharmacological treatment. Non-inclusion criteria were neurological disorders or sequelae other than neuropathic pain; psychiatric disorders other than anxiety or depressive comorbidities; a history of head injury with loss of consciousness or stroke; belonging to a protected population (e.g. pregnant women or individuals under legal guardianship); inability to provide informed consent; and severe uncorrected hearing or visual impairment that prevented completion of the assessment.

In total, 106 participants completed the study. They were excluded from analyses (n=5) in cases of incomplete assessments, missing data, or non-compliant behavior during testing sessions.

Participants received financial compensation of €50 upon completion of both the inclusion and experimental sessions. All participants provided written informed consent prior to participation. The study was approved by the local Ethics Committee (CER Unistra: 2022-44) and was conducted in accordance with the Declaration of Helsinki.

### 2.2. Study design and protocol

Each participant was assigned a unique identifier and password, allowing pseudonymous authentication on the application. Standardized instructions on tablet use were provided prior to testing. Testing sessions were conducted in the presence of an experimenter, who remained in the background to address potential technical issues (e.g. device malfunction) but did not interfere with task performance. After receiving initial instructions, participants completed the assessment autonomously, as the tool was specifically designed for self-administration without examiner input.

The experimental design (Figure 1) implemented to assess the different validation criteria was adapted from previous methodological frameworks (Laforce et al., 2018).

**Figure 1:**
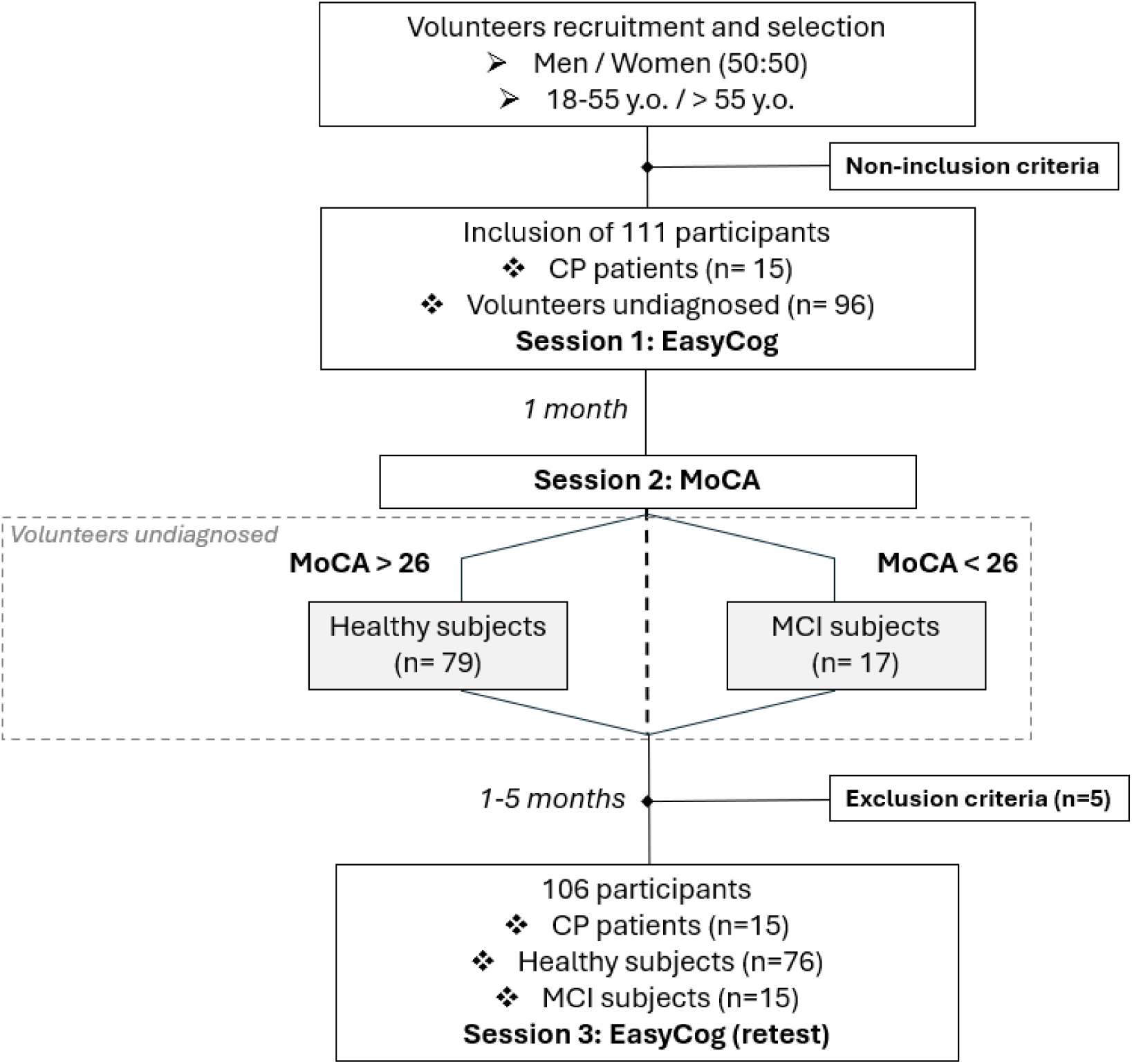
Flowchart of the study design. The validation study of the EasyCog tool began by recruiting 111 undiagnosed volunteers. Non-inclusion criteria were checked during the interview in session 1, before the first EasyCog evaluation. One month later, all participants took part in session 2, in which they were divided into healthy subjects (n=79) and MCI subjects (n=17) based on the MoCA score (threshold = 26). From 1 to 5 months later, 96 participants completed the study, with session 3 designed to assess the tool’s test-retest effect. Five participants were *a posteriori* excluded for non-full completion of the evaluations, missing data, or unserious behavior during sessions. CP patients (n= 15) followed the same study design. MCI: mild cognitive impairment; CP: chronic pain.

All participants first completed an initial session using the EasyCog tool (session 1), which contributed to the standardization process by ensuring consistent administration conditions (e.g. instructions, task order).

One month later, participants completed the French paper version of the Montreal Cognitive Assessment (MoCA, version 8.3; session 2). This session served both to classify participants into two groups: healthy subjects (HS; MoCA ≥ 26; n = 79) and individuals with mild cognitive impairment (MCI; MoCA < 26; n = 17); and to examine convergent validity by comparing EasyCog performance with a validated reference measure.

Finally, participants were retested with the EasyCog tool 1 to 5 months after their second session (session 3) to assess test-retest reliability across different time intervals.

Participants with chronic pain (CP) underwent the same protocol, which was integrated into their usual care pathway. Their inclusion allowed the assessment of EasyCog specificity through comparisons with both MCI and healthy groups.

### 2.3. Material and methods

#### Digital tool and artificial intelligence

EasyCog was developed by the team in collaboration with Traboule Labs (Strasbourg, France). This Android tablet application offers a battery of tests (MoCA-like and MMSE-like) and enables self-assessment of cognitive abilities through a speaking character (human-like voice) that interacts with the subject. Among the aimed users, the senior population has a different culture regarding tactile and digital interfaces, when it has one at all, compared to the younger population. The personification allows for a better understanding and involvement of these patients in the test (Lu et al., 2017).

Verbal answer transcriptions are ensured by a serverless artificial neural network (VOSK, Kaldi) embedded into the application. The pseudonymized results were directly transmitted to a secure server, where they could be downloaded as an Excel file.

#### Self-assessment of fatigue, stress, and pain

Several parameters have been integrated into the tool to assess the key variables that influence cognitive abilities, including fatigue, stress, and pain. These latter parameters were assessed using intensity and unpleasantness visual analog scales (VAS), ranging from 0 (none) to 100 (maximum). These allow participants to make a self-assessment based on their feelings, as they lack numerical indicators (Jamison et al., 2002).

#### Cognitive self-assessment and scoring

The different tests integrated into the application assess several cognitive domains, including attention, memory, executive functions, language, visuospatial abilities, and constructional skills. For all tests images, numbers, and words were randomized to minimize learning effects during repeated assessments. For the Bell test, sticker positions were randomized according to the rules described in the original paper (Gauthier et al., 1989).

The 11 cognitive tests included in EasyCog were adapted from the validated existing MoCA (Rossetti et al., 2011) and MMSE (Crum et al., 1993) batteries (i.e., MoCA-like and MMSE-like tests), excluding the bell test.

These tests were scored either according to the MoCA system (i.e., EasyCog MoCA scores) or the MMSE system (i.e., EasyCog MMSE scores), depending on which scoring method allowed for a broader distribution of points. Consequently, the MMSE-like and MoCA-like tests included in EasyCog assess 23/30 and 24/30 points of the original batteries, respectively (Table 1). Rather than relying solely on cut-off scores, we analyzed performance dimensionally by grouping task scores according to the cognitive functions they engaged. Domain-specific scores were then computed and expressed as percentages of the maximum obtainable score, thereby enabling the establishment of cognitive profiles visualized with spider plots (see Figure 4).

**Table 1:**
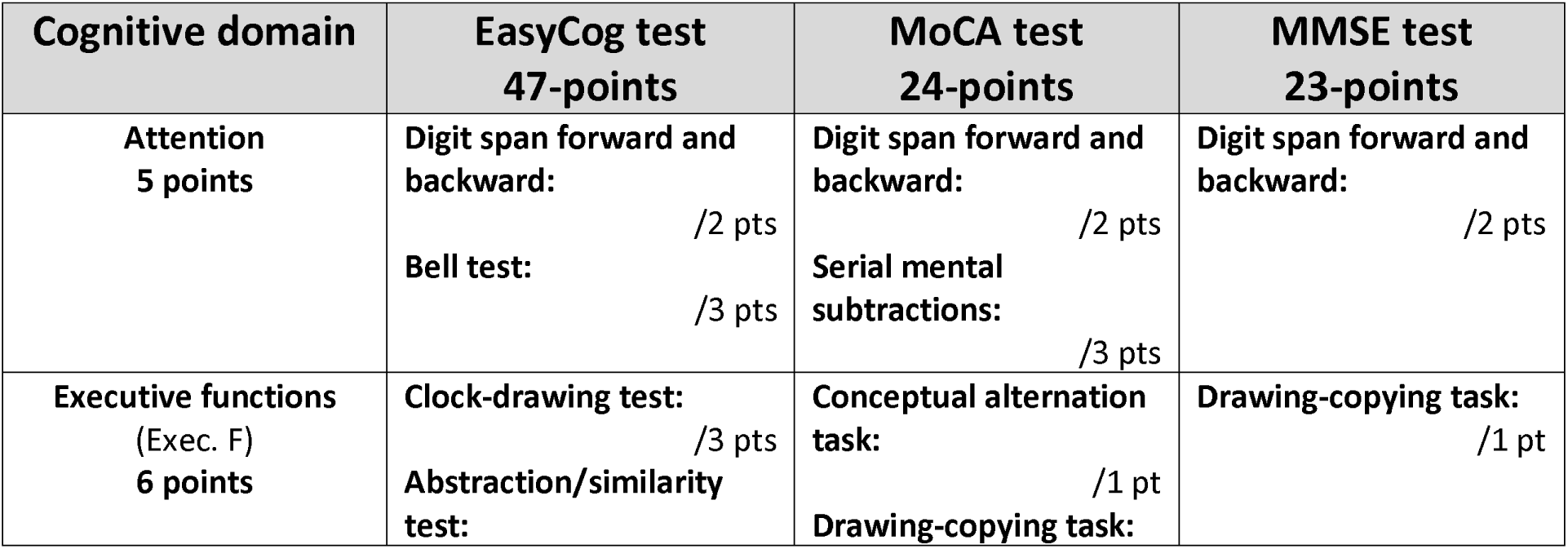
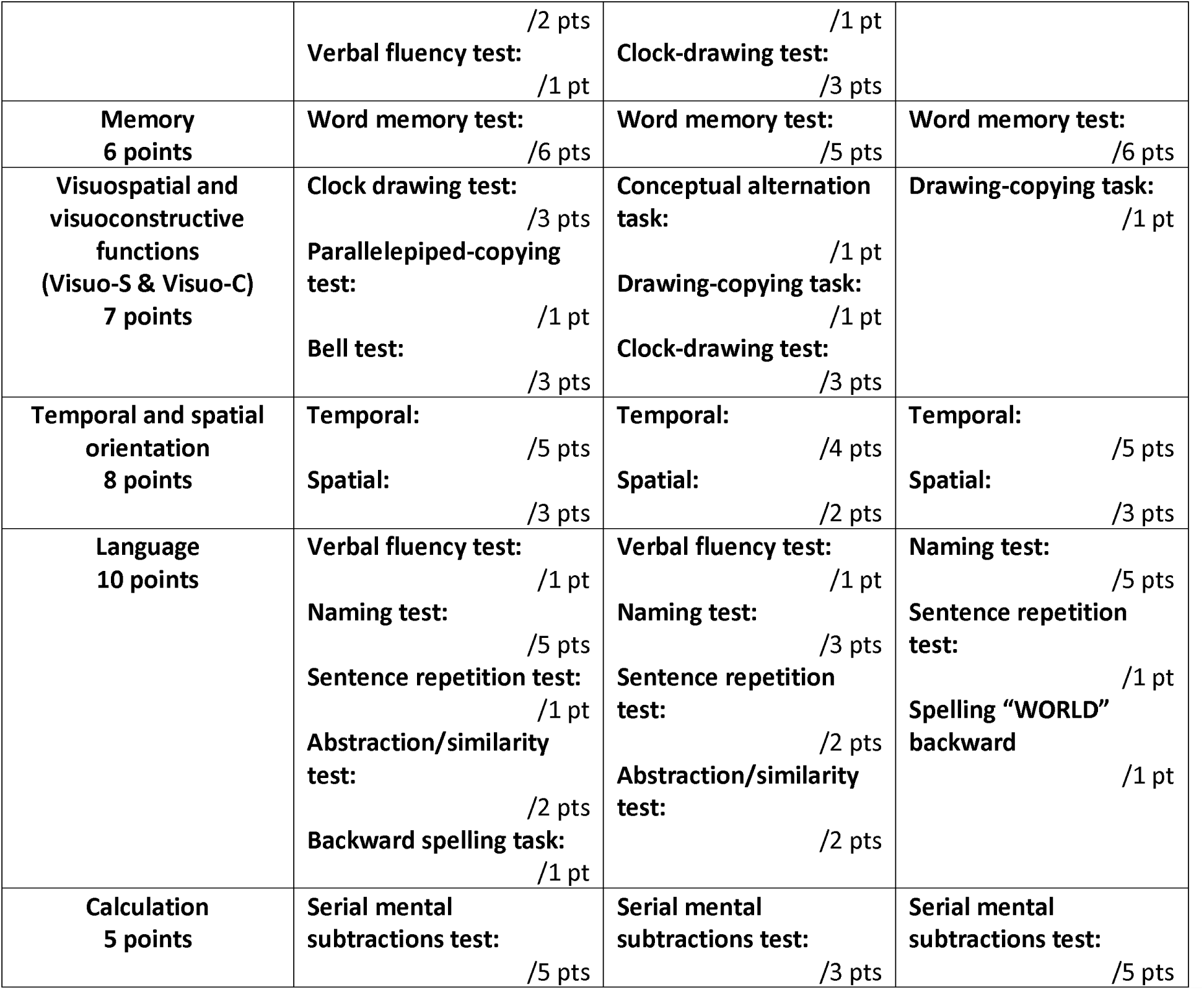
Cognitive functions assessed and comparative scoring of EasyCog, MoCA and MMSE. Each cognitive function is assessed by several tests/questions associated with a score. Total MoCA and MMSE scores are out of 30 points. EasyCog offers a 47-point evaluation. Of this total score, 23 and 24 points from the MMSE and MoCA tests, respectively, represent 41 points of the EasyCog score because of redundancy between the MMSE and MoCA. A modified bell test score (out of 3 points) is incorporated into scores for attention and visuospatial and visuoconstructive abilities.

##### Temporal and spatial orientation tests

Participants are asked a series of questions assessing their orientation in time and space (e.g., date, location). This task evaluates orientation abilities, which rely on memory and general cognitive functioning (Folstein et al., 1975; Nasreddine et al., 2005).

##### Word memory test

A list of three words is orally presented to participants at a rate of one word per second. Participants are first asked to recall the words immediately. After a delay, during which they perform other tasks, they are asked to recall the words again. This task assesses verbal memory, including immediate recall and delayed retrieval processes (Benton & Hamsher, 1978; Folstein et al., 1975).

##### Naming test

Participants are presented with drawings of three animals and two objects and are asked to name them. This task assesses language abilities, particularly lexical access, as well as visual recognition processes (Kaplan, 1983).

##### Digit span forward and backward

The numerical span task consists of two sequences of digits (five and three digits). In the first condition, participants repeat the digits in the same order, whereas in the second condition, they repeat them in reverse order. This task assesses attention and working memory, with the backward condition placing additional demands on executive processes such as information manipulation and updating (Baddeley, 2003; Miller, 1956).

##### Mental subtraction test

Participants are required to perform five consecutive mental subtractions of seven starting from 100. This task assesses calculation abilities and engages attention and executive functions, particularly working memory and cognitive load (Karzmark, 2000; Nasreddine et al., 2005).

##### Abstraction/similarity test

Participants are asked to identify a conceptual similarity between two words (e.g. orange and banana: fruits). This task assesses abstract reasoning and semantic processing, reflecting higher-level conceptualization abilities (Nasreddine et al., 2005; Raoux et al., 2014).

##### Clock-drawing test

This test involves drawing a clock, placing all the digits, and indicating a given hour. Drawing the clock frame requires visuoconstructive skills. Placing and drawing the numbers use the language function of numbers, as well as visuospatial skills. Finally, placing and drawing the clock hands tests executive abilities (Budson & Solomon, 2016).

##### Verbal fluency test

Participants are asked to generate as many words as possible within one minute, beginning with a letter randomly selected from among those that are the initial letter of at least twenty French words. Proper nouns, numbers, and variations of the same word are excluded. This task assesses phonemic verbal fluency and relies on executive processes such as strategic search and cognitive flexibility (Benton & Hamsher, 1978).

##### Sentence repetition test

Participants are asked to repeat one sentence verbatim. This task assesses language abilities, including phonological processing and syntactic integration, and is sensitive to impairments in speech production, as observed in aphasic disorders (Bourgeois-Marcotte et al., 2015).

##### Backward spelling task

Participants are asked to spell a word backward, thereby assessing spelling abilities while also engaging attention and working memory.

##### Parallelepiped-copying test

Participants are asked to reproduce a drawing of a parallelepiped as accurately as possible. This task primarily assesses visuoconstructive abilities and visuospatial processing (Nasreddine et al., 2005).

##### Bell test

Participants are asked to circle 35 target bells embedded among 280 distractors (e.g., houses, horses), arranged across seven columns. This task assesses selective and sustained attention, as well as visuospatial exploration. Patterns of omissions may indicate attention deficits or unilateral spatial neglect, particularly when omissions are lateralized (Gauthier et al., 1989).

### 2.4. Statistics

Data normality was assessed using the Shapiro–Wilk test. For normally distributed variables, group comparisons (including sex differences) were performed using a two-way ANOVA. When normality assumptions were not met, non-parametric tests were used. Specifically, one-sample Wilcoxon signed-rank tests were conducted to compare observed medians against reference values, Mann–Whitney U tests were used for between-group comparisons, and Kruskal–Wallis tests were applied for comparisons involving more than two groups. When the Kruskal–Wallis test revealed a significant effect, post hoc multiple comparisons were performed using Dunn’s test with adjusted p-values. All statistical analyses were conducted using GraphPad Prism (version 6.0).

## 3. Results

### 3.1. Sociodemographic and Visual Analog Scale (VAS) data

Seventy-six healthy subjects (HS; mean age = 48.6 ± 20.2 y/o) were included in the analysis with equal representation of males (n= 38, mean age = 49.3 ± 21.0 y/o) and females (n= 38, mean age = 49.6 ± 21.5 y/o). The demographic data for HS included in this work are presented by age range in Table 2, along with VAS values for fatigue, stress, and pain. The subjects are balanced by sex and well distributed by age (18 to 93 y/o). However, there was a greater prevalence of high educational levels (ranging from 0 to 4), which were generally higher than 12 years of education, but equivalent across sex (females mean = 3.2 ± 1.0; males mean = 3.4 ± 0.4) and age ranges (except for females over 75 y/o). Mean VAS scores are low, as expected, for fatigue (mean= 18.8 ± 19.1), for stress (mean= 10.6 ±18.4), and for pain (mean= 6.6 ± 14.8). No significant interaction was observed between VAS dimensions (intensity vs unpleasantness ratings) and age group, sex, or educational level. However, significant main effects were found for age (ANOVA: F_(6,300)_ = 3.15, p = 0.005) and educational level (ANOVA: F_(3,318)_ = 2.91, p = 0.03), whereas no significant main effect of sex was observed (ANOVA: F_(1,342)_ = 0.19, p = 0.66).

**Table 2:** Sociodemographic and Visual Analog Scale (VAS) data from Healthy Subjects (HS). Of the healthy subjects included (38 male and 38 female), the age distribution ranged from 18 to 93, and the level of education from 1 to 4 (0: no diploma; 1: below 9 years of study; 2: below 11 years of study; 3: A level / 12 years of study; 4: higher education). The visual analog scales (VAS) evaluated during the EasyCog sessions confirmed that healthy subjects’ scores remained low overall. Statistical analysis using a 2-way ANOVA revealed no effect of sex or education level on mean VAS scores, for the intensity (int.) or the unpleasantness criterion (unp.).

|  |  | **18-25** | **25-35** | **35-45** | **45-55** | **55-65** | **65-75** | **>75** | **TOTALS** |
| --- | --- | --- | --- | --- | --- | --- | --- | --- | --- |
| **N** |  | **15** | **8** | **11** | **9** | **10** | **16** | **7** | **76** |
| **Sex** | F | 9 | 4 | 6 | 6 | 4 | 7 | 2 | **38** |
|  | M | 6 | 4 | 5 | 3 | 6 | 9 | 5 | **38** |
| **Age**<br>(mean ± SD) | F | <u>21,3</u><br>$\pm 2,4$ | <u>27,3</u><br>$\pm 2,6$ | <u>38,8</u><br>$\pm 2,5$ | <u>49,7</u><br>$\pm 2,9$ | <u>63,0</u><br>$\pm 1,4$ | <u>69,7</u><br>$\pm 1,5$ | <u>77,5</u><br>$\pm 0,7$ | <u>49,6</u><br>$\pm 21,5$ |
| | M | <u>22,2</u><br>$\pm 1,3$ | <u>29,3</u><br>$\pm 2,2$ | <u>38,8</u><br>$\pm 1,1$ | <u>47,0</u><br>$\pm 2,0$ | <u>59,7</u><br>$\pm 3,2$ | <u>70,1</u><br>$\pm 3,4$ | <u>78,4</u><br>$\pm 3,1$ | <u>49,3</u><br>$\pm 21,0$ |
| **Educational** | F | <u>3,4</u> | <u>4,0</u> | <u>3,7</u> | <u>3,8</u> | <u>2,8</u> | <u>3,4</u> | <u>1,0</u> | <u>3,2</u> |
| **level** | | $\pm 1,0$ | $\pm 0,0$ | $\pm 0,5$ | $\pm 0,4$ | $\pm 1,0$ | $\pm 0,8$ | $\pm 0,0$ | $\pm 1,0$ |
| (mean ±SD) | M | <u>3,5</u><br>$\pm 0,8$ | <u>4,0</u><br>$\pm 0,0$ | <u>3,7</u><br>$\pm 0,4$ | <u>3,5</u><br>$\pm 0,4$ | <u>3,4</u><br>$\pm 0,8$ | <u>2,7</u><br>$\pm 1,4$ | <u>3,0</u><br>$\pm 1,0$ | <u>3,4</u><br>$\pm 0,4$ |
| **Fatigue VAS** | Int | <u>27,6</u><br>$\pm 14,4$ | <u>34,4</u><br>$\pm 12,3$ | <u>33,9</u><br>$\pm 23,5$ | <u>28,9</u><br>$\pm 24,0$ | <u>20,2</u><br>$\pm 18,2$ | <u>18,1</u><br>$\pm 24,7$ | <u>17,0</u><br>$\pm 17,4$ | **18,8**<br>$\pm 19,1$ |
| (mean ±SD) | Unp | <u>14,1</u><br>$\pm 13,2$ | <u>4,6</u><br>$\pm 6,7$ | <u>17,1</u><br>$\pm 22,9$ | <u>14,3</u><br>$\pm 15,0$ | <u>8,7</u><br>$\pm 14,0$ | <u>10,4</u><br>$\pm 13,8$ | <u>5,0</u><br>$\pm 10,8$ | |
| **Stress VAS** | Int | <u>8,1</u><br>$\pm 11,6$ | <u>14,8</u><br>$\pm 21,6$ | <u>18,1</u><br>$\pm 27,5$ | <u>6,1</u><br>$\pm 10,2$ | <u>16,1</u><br>$\pm 22,0$ | <u>8,9</u><br>$\pm 17,3$ | <u>11,9</u><br>$\pm 20,5$ | **10,6**<br>$\pm 18,4$ |
| (mean ±SD) | Unp | <u>5,5</u><br>$\pm 6,7$ | <u>10,5</u><br>$\pm 18,8$ | <u>13,1</u><br>$\pm 22,0$ | <u>16,2</u><br>$\pm 33,0$ | <u>11,0</u><br>$\pm 18,3$ | <u>8,2</u><br>$\pm 15,3$ | <u>7,4</u><br>$\pm 10,4$ | |
| **Pain VAS** | Int | <u>4,9</u><br>$\pm 10,7$ | <u>5,0</u><br>$\pm 14,1$ | <u>5,1</u><br>$\pm 16,9$ | <u>1,8</u><br>$\pm 3,6$ | <u>8,5</u><br>$\pm 11,1$ | <u>9,4</u><br>$\pm 20,6$ | <u>6,1</u><br>$\pm 11,1$ | **6,6**<br>$\pm 14,8$ |
| (mean ±SD) | Unp | <u>7,6</u><br>$\pm 16,7$ | <u>4,9</u><br>$\pm 13,8$ | <u>0,1</u><br>$\pm 0,3$ | <u>0,0</u><br>$\pm 0,0$ | <u>10,0</u><br>$\pm 15,2$ | <u>12,6</u><br>$\pm 24,3$ | <u>8,6</u><br>$\pm 10,9$ | |

The demographic data of MCI participants (**see Table S1, Supplementary data**) show a higher prevalence among males (11 males and 4 females), and the mean age was 67.5 ± 13.8 y/o. Their educational level was lower than that of the HS group (mean = 2.7 ± 1.1). Their VAS scores were similar to the HS group (fatigue mean = 16.1 ± 22.9; stress mean = 5.5 ± 9.8; pain mean = 9.9 ± 16.2).

While CP patients’ data (**see Table S2, Supplementary data**) showed an equivalent proportion of males and females (8 males and 7 females) and a mean age of 68.3 ± 9.0 y/o. Their educational level was similar to that of the MCI group and lower than that of the HS participants (mean = 2.3 ± 1.0). Their clinical profile was highly variable (e.g. musculoskeletal pain, chronic low back pain, neuropathic pain, rheumatoid arthritis, fibromyalgia, chronic migraine…) and their treatments as well (e.g. opioids, benzodiazepines, antidepressants, antiepileptics…etc.). The VAS score of CP patients was logically higher than the other groups (fatigue mean = 36.3 ± 29.3; stress mean = 26.1 ± 31.0; pain mean = 38.2 ± 37.0). Interestingly, males subjectively rated their pain lower than females on both intensity and unpleasantness criteria.

### 3.2. Voice recognition performance

The innovation of EasyCog relies on the subjects’ autonomy during the tests and, therefore, on the performance (both accuracy and processing speed) of the voice recognition provided by the artificial neural network (ANN) embedded in the digital tablet. Accuracy is mandatory for automated analysis, while processing speed on the device affects the user experience and, in turn, their involvement in the procedure. Regarding the latter point, in our setting, the maximum processing time was 10 seconds for 1 minute of voice signal – an extreme case. In most cases, waiting/loading times were never more than 2 or 3 seconds between questions. On the accuracy side, shortcomings were observed in questions involving: a proper noun unknown to the ANN (e.g., a small village name), repetition of a “wrong” sentence (phonetic one, without any proper syntax or grammar), and the naming of animals/objects without context which have homonyms (e.g., the French “paon” vs “pont”). For this validation work, incorrectly transmitted answers were corrected from notes taken during sessions for each subject.

### 3.3. Reliability: EasyCog as an alternative for MoCA and MMSE evaluation

We first compared the normative values of MoCA and MMSE (paper versions; data available in the literature and reference websites) with EasyCog-MoCA and EasyCog-MMSE scores, normalized at 30. As for MoCA and MMSE, there was a slight reduction in the values with age.

Across all ages, no statistical differences were found between EasyCog-MoCA scores and normative MoCA values reported in the literature (2-way ANOVA: interaction group x age, F_(1,2883)_=1.537; p=0.21; Figure 2A). Contrary to MoCA, a statistical difference is observed between MMSE normative values and EasyCog-MMSE (2-way ANOVA: interaction group x age, F_(1,17755)_=3.859; p=0.0008). Post hoc analysis indicated that this was due only to the extreme age (18-25 and >75 y/o), displaying lower mean values (Figure 2B).

**Figure 2:**
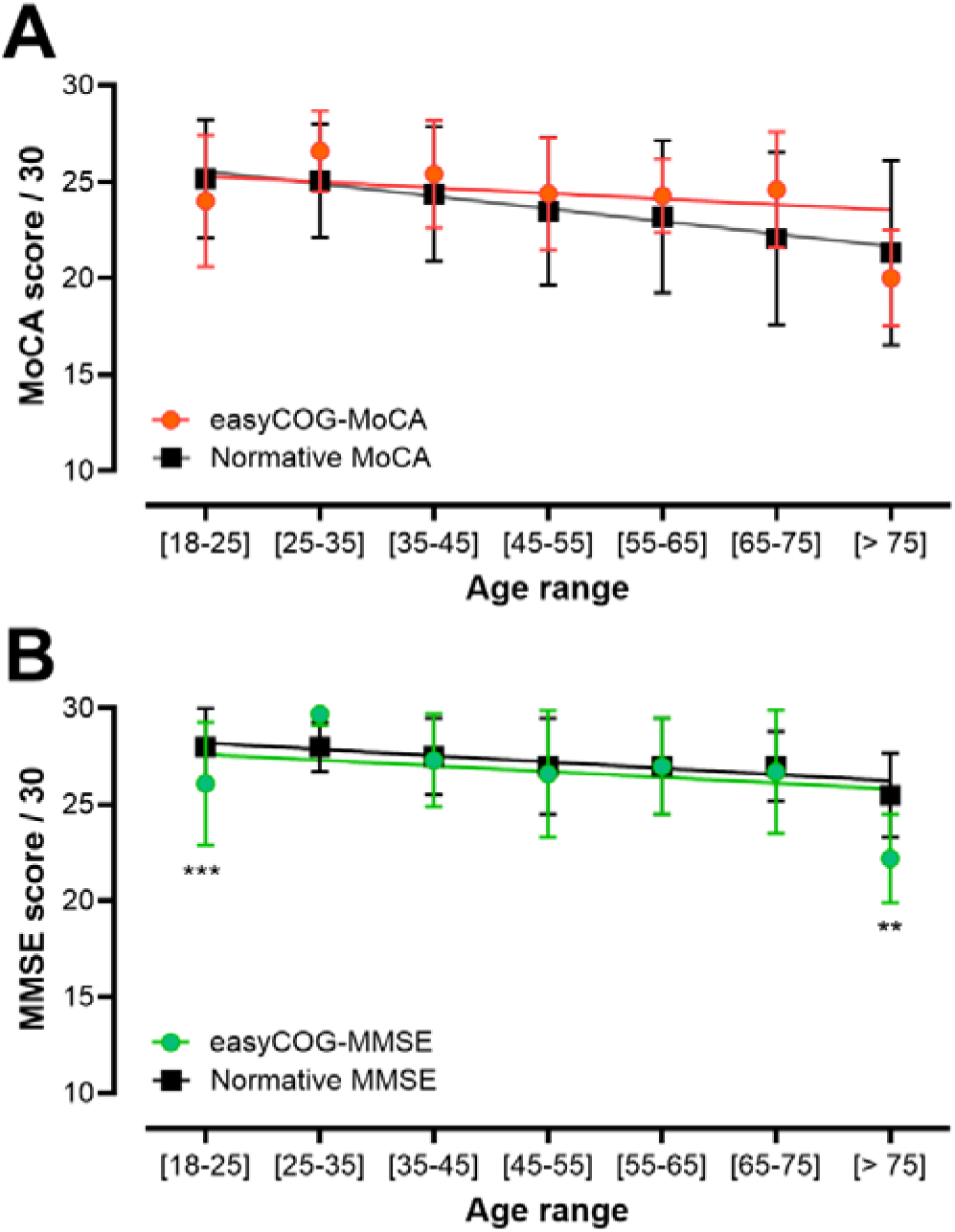
EasyCog MoCA **(A)** and EasyCog MMSE **(B)** scores fitting with normative values. After normalization to 30 points, scores are expressed as mean ± SD for each age range. Mean scores of EasyCog MoCA and MMSE exhibit a similar reduction across age ranges, consistent with the normative values for MoCA **(A)** and MMSE **(B)**. Statistical code: Tukey’s multiple comparison post hoc test at p<0.01 (**) and p<0.001 (***).

### 3.4. Scoring cognitive functions with EasyCog

Panels in Figure 3 shows the breakdown of EasyCog cognitive functions (score expressed as a percentage of the maximal possible performance). It nicely shows the age-dependent reduction in some cognitive functions, such as executive functions, visuospatial and visuoconstructive functions, and memory. When looking at the corresponding histograms, scores of two functions, i.e., executive function (χ^2^(2) =6.29; p=0.04), visuospatial and visuoconstructive functions (χ^2^(2) =10.16; p=0.006) are significantly lower for the older age group (>65 yo) when compared to the younger age group (see Figure 3). However, histograms from other cognitive functions assessed were not different with age (i.e. calculation, χ^2^(2) =2.17; p=0.34), attention (χ^2^(2) =0.33; p=0.85), language (χ^2^(2) =2.11; p=0.35), orientation (χ^2^(2) =2.78; p=0.25), despite a tendency for memory (χ^2^(2) = 5.43; p=0.06). Nevertheless, when examining the mean EasyCog global scores (i.e., the mean of all function scores), a significant difference between the older and younger age groups was again observed, indicating global age-related cognitive decline (χ^2^(2) =9.51; p=0.008; Figure 3).

**Figure 3:**
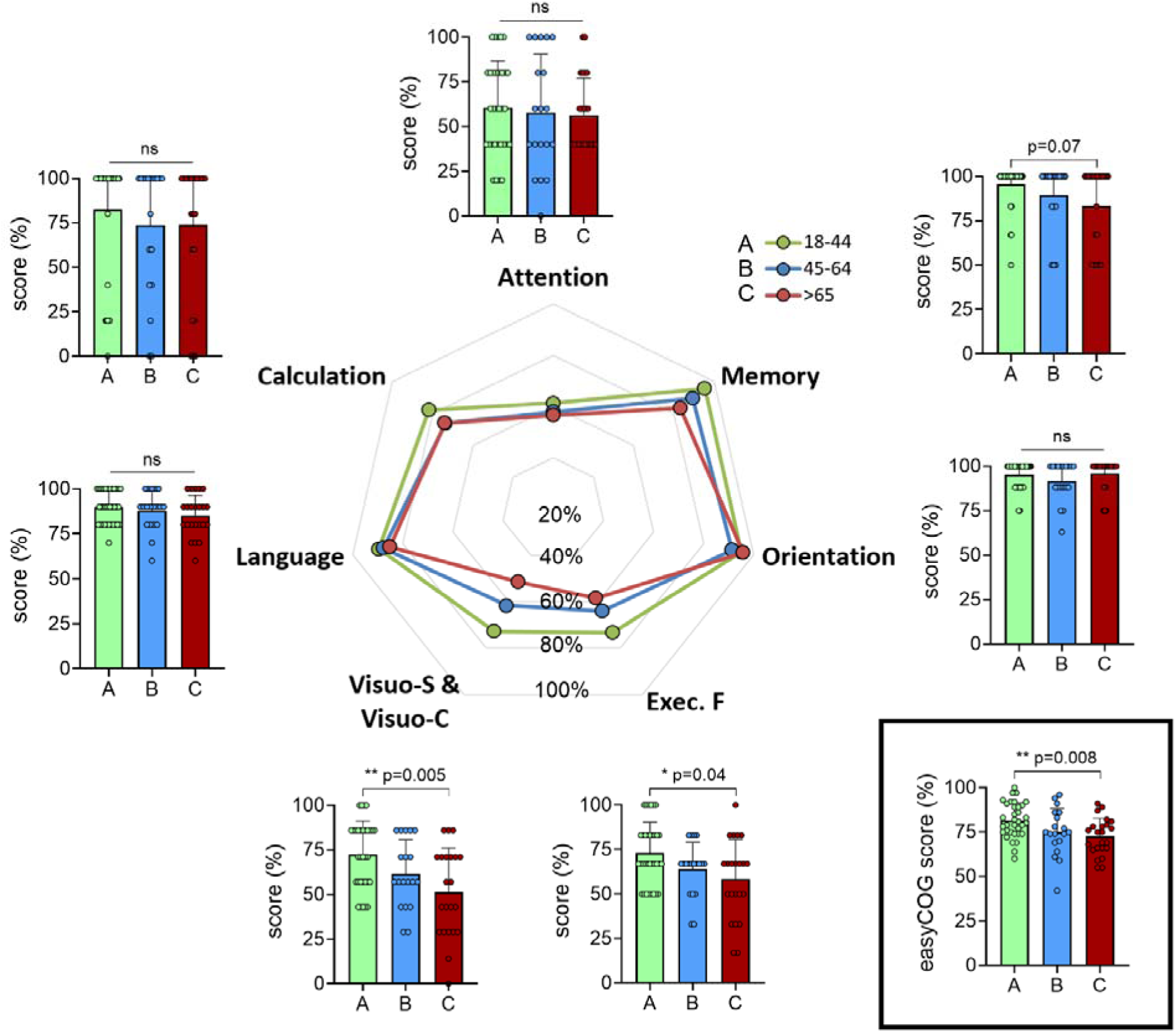
Breakdown of EasyCog scores per cognitive function. The spider plot in the middle illustrates changes in the mean EasyCog scores for the indicated cognitive functions across age classes. Histograms are also provided for each cognitive function and global EasyCog score, with mean ± SD values. Note that age-dependent significant changes could be observed for some functions: executive function and visuospatial and visuoconstructive functions; and for the EasyCog global score (general statistics, Kruskal-Wallis). Statistical code (*) and p values correspond to Dunn’s post hoc test.

### 3.5. Test-retest reliability

The test-retest reliability results are shown in Figure 4. The mean differences (deltas) between test and retest scores for HS were calculated and expressed as percentages. Wilcoxon Signed Rank statistical analyses revealed no significant difference from 0 for memory (W= −2, p > 0.99; W= 2, p= 0.94; W= −12, p=0.19), orientation (W= −15, p= 0.34; W= −7, p= 0.79; W= −14, p=0.36), language (W= 16, p=0.42; W= 5, p > 0.99; W= 39, p=0.06), and calculation (W= 1, p= 0.99; W= 16, p= 0.17; W= 3, p=0.91) for all delays respectively (i.e., 2 months, 3 months, > 4 months). For attention, no significant difference from 0 was found at a delay of 3 months (W = 55, p = 0.09). Deltas from both executive functions (3-months: W = 48, p = 0.06; > 4-months: W = 10, p = 0.65) and visuospatial and visuoconstructive functions (3-months: W = 59, p = 0.07; > 4-months: W = 18, p = 0.42) showed no significant difference from 0 at 3-months and >4-months delays. Finally, statistical analyses revealed a significant difference from 0 for deltas of EasyCog global scores for a 2-month (W= 99, p= 0.02) and 3-month (W= 127, p= 0.01) delay, but no significant difference from 0 for the >4-month delay (W= 53, p= 0.22).

**Figure 4:**
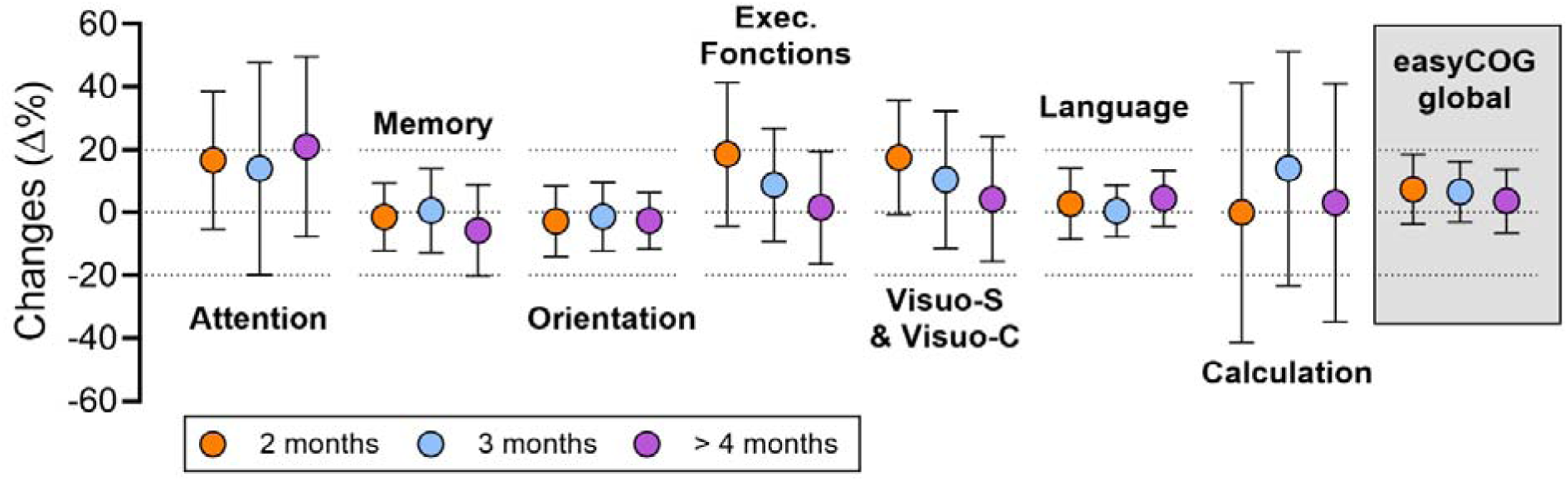
Test-retest reliability of EasyCog. Deltas between scores of the two EasyCog sessions were calculated and expressed as a percentage (i.e., changes Δ%). The delay between the two EasyCog sessions was either 2 months, 3 months, or greater than 4 months but less than 6 months (i.e., > 4 months). Wilcoxon Signed-Rank statistical analyses revealed no significant difference from 0 for deltas of all functions and EasyCog global scores at >4 months’ delay.

### 3.6. Specificity and sensitivity

Among the volunteers, some subjects exhibited a MoCA score < 26 (paper version) and were classified into the MCI group (see the average MoCA score in Figure 5). Their MoCA (Mann-Whitney U = 0, p < 0.001) and EasyCog global (Mann-Whitney U = 267, p < 0.001) scores were both significantly lower than HS’s values for the same age group. As for MCI subjects, CPP showed significantly lower MoCA (Mann-Whitney U = 28, p < 0.001) and EasyCog scores (Mann-Whitney U = 244.5, p < 0.001) than HS. Interestingly, although no differences were observed between the averaged scores of MCI and CPP, the EasyCog profiles differed strikingly. Indeed, when comparing EasyCog scores for each cognitive function tested across the MCI group, CPP, and HS, cognitive profiles differed substantially (Figure 5, spider plots). Compared to MCI, CPP had reduced mean scores for attention (CPP: 40.0±21%; MCI: 46.3±27%) and calculation performance (CPP: 58.5±40%; MCI: 65.0±38.0%), visuospatial and visuoconstructive functions (CPP: 33.7±24%; MCI: 63.4±21%), and poorer memory (CPP: 74.4±26.0%; MCI: 82.3±28.0%). Under these conditions and with respect to the large variability observed, only visuospatial and visuoconstructive functions were significantly different between CPP and MCI subjects (Mann Whitney U=40, p=0.002). This highlighted interest in using the EasyCog profile to finely evaluate specific deficits, monitor disease progression or the treatment’s adverse/positive effects, and possibly address them with appropriate care.

**Figure 5:**
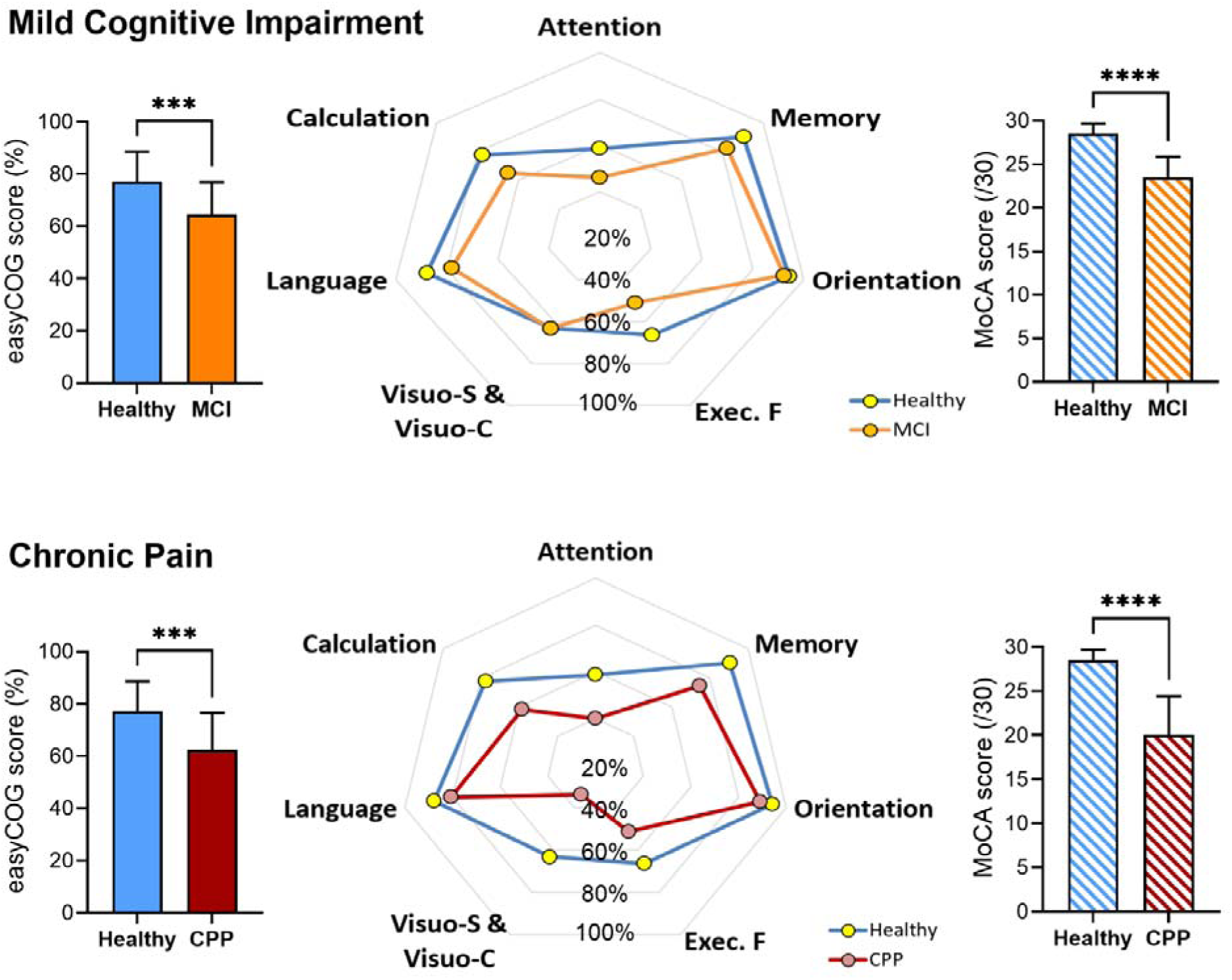
EasyCog sensitivity to different cognitive impairments. Histograms show that both MCI and CPP have significantly lower MoCA and EasyCog global scores than HS (Mann-Whitney, *** p < 0.001 and **** p<0.0001). The spider plots in the middle illustrate changes in the EasyCog mean scores for the indicated cognitive functions in two distinct populations with different cognitive impairments (MCI at the top and CPP at the bottom) compared to the HS. Note that CPPs have even lower EasyCog scores than MCI for some functions (i.e., attention, calculation, memory, visuospatial, and visuoconstructive functions).

## 4. Discussion

The present study aimed to validate EasyCog, a novel digital self-administered cognitive assessment tool designed to provide a global evaluation of cognitive functions while remaining suitable for routine clinical use. Overall, the results support the tool’s validity, reliability, sensitivity, specificity, and test-retest reproducibility. In addition, EasyCog enabled the characterization of distinct cognitive profiles in healthy individuals, subjects with mild cognitive impairment (MCI), and patients with chronic pain (CP).

The demographic characteristics of the healthy sample were reassuringly homogeneous for the population included in this validation study. We observed no interaction between sociodemographic variables and visual analog scale (VAS) ratings for fatigue, stress, and pain, suggesting that these factors did not primarily explain differences in cognitive performance. Most participants had a relatively high level of education, which was well balanced across age groups and between sexes. This point remains important, as educational level is known to influence performance on cognitive screening tests (Oz et al., 2022; Ramos-Henderson et al., 2025) and may contribute to cognitive reserve, which delays cognitive decline (Le Carret et al., 2003; Stern, 2002; Tucker-Drob et al., 2009). Because MCI and CP participants had a higher mean age than HS, we also compared them with an age-matched subgroup of HS and found no evidence that age in the MCI or CP group influenced the effect on altered functions.

EasyCog scores in healthy participants showed an age-related cognitive decline consistent with the literature on aging (Haaland et al., 2003; Klimova, 2025). Once normalized, EasyCog-MoCA and EasyCog-MMSE scores followed regression curves similar to those reported for paper versions of the MoCA and MMSE, supporting the tool’s convergent validity. However, a slight discrepancy was observed in the youngest and oldest age groups compared with normative MMSE values reported in the literature (Crum et al., 1993). This statistical difference likely results from the relatively limited sample size in these extreme age categories compared with the large normative cohorts available (>11000 subject values).

At the functional level, EasyCog detected age-related declines in executive and memory functions, which are among the cognitive domains most consistently affected during normal aging (Balota et al., 2000; Bisiacchi et al., 2008; Lu et al., 2016; Rönnlund et al., 2005). These findings are consistent with neuroimaging studies reporting preferential age-related atrophy and reduced connectivity in frontal brain regions, along with associated executive dysfunction (Balota et al., 2000; Bisiacchi et al., 2008; Lu et al., 2016; Rönnlund et al., 2005). Interestingly, visuospatial and visuoconstructive functions also appeared sensitive to aging in our sample, consistent with previous reports of age-related alterations in parietal and occipital networks involved in spatial processing (Wiesman & Wilson, 2019).

In contrast, no significant age effect was observed for attentional scores. This result was unexpected, given that attentional and executive processes are commonly considered among the earliest functions affected by aging (Kaufman et al., 2016; Lu et al., 2016). EasyCog results revealed substantial variability in attentional performance within each age group, as well as relatively low mean attentional scores, even among younger adults aged 18–25 years. Surprisingly, this youngest age group even showed lower mean attentional scores than participants aged 35–45 years. Although speculative, this observation raises questions about the potential influence of modern lifestyles on attentional capacities, particularly among younger generations exposed to rapid information flow, multitasking environments, and short-form digital content (Firth et al., 2019; Mittal et al., 2026). These findings should nevertheless be interpreted cautiously due to the inter-individual variability, the lack on age-related linearity of the attentional scores and, across age, the average low scores seen in healthy subjects (around 60%) that reflect the demanding nature of the tasks which strongly engages sustained and selective attention resources.

An important contribution of EasyCog lies in its dimensional approach to cognitive assessment. Rather than relying exclusively on cut-off scores, task performances were grouped according to the cognitive functions involved, allowing the computation of domain-specific scores expressed as performance percentages. This approach enabled the visualization of individualized cognitive profiles through spider plots and provided a more detailed characterization of cognitive functioning than global scores alone. Importantly, this method highlighted distinct cognitive patterns between MCI and CP participants despite similarly reduced global cognitive scores.

Although cognitive impacts of CP are frequently compared with MCI (Corti et al., 2021; Patel et al., 2025), both associated with increased risk of dementia (Kumaradev et al., 2021; Whitlock et al., 2017), our results suggest that CP states are associated with specific cognitive alterations. This aligns with previously published studies (Liu et al., 2014; Moriarty et al., 2011; Oosterman et al., 2012). Several mechanisms may explain this specificity. For instance, CP likely captures attentional resources through excessive nociceptive processing within the cognitive components of the pain neuromatrix, thereby interfering with ongoing cognitive tasks (Eccleston et al., 1997; Grisart & Van der Linden, 2001). Executive dysfunction may also be reinforced by emotional dysregulation resulting from alterations in mesocorticolimbic pathways involved in motivation and reward processing, as well as hyperactivity of the anterior cingulate cortex associated with anxiodepressive symptoms (Caes et al., 2021; Letzen et al., 2019; Moriarty et al., 2011; Solberg Nes et al., 2009). Memory dysfunction may also relate to maladaptive neuroplasticity (Moriarty et al., 2011), neuroinflammatory processes that affect prefrontal and hippocampal function (Li et al., 2025), and impaired extinction of aversive memories (Dunsmoor et al., 2015; Stegemann et al., 2023).

In this context, identifying specific vulnerable cognitive domains appears clinically relevant for managing CP patients. EasyCog reliability showed satisfactory reproducibility after sufficiently spaced retest intervals. Randomizing images, words, numbers, and letters across sessions limited learning effects for most cognitive domains, allowing longitudinal follow-up. Indeed, no significant retest effect was observed after two months for most functions. For attentional, executive, visuospatial, and visuoconstructive functions, learning effects disappeared after three months. Because EasyCog global score still showed residual effects at this interval, a minimum retest interval of four months appears preferable to ensure optimal reproducibility with time.

Beyond its psychometric properties, EasyCog has several advantages for clinical use and research. Embedded artificial intelligence-based voice recognition enables self-administration without an examiner’s intervention. This reduces inter-rater variability and facilitates its implementation in hospital settings, primary care, or remote assessment contexts. Furthermore, the digital format enables the integration of additional biomarkers such as reaction times and automated analysis of task performance, which may further improve EasyCog sensitivity.

Several limitations should nevertheless be acknowledged. First, the sample size of this study remained relatively limited, particularly for extreme age groups and clinical subgroups, which may have reduced statistical power for secondary factor analyses (sex specificity, education, etc.). Second, most healthy subjects have relatively high educational levels, potentially overestimating the current normative data. To address these points, we plan to continue testing healthy subjects from diverse origins and socio-economic backgrounds to enrich the database. Third, we classified MCI participants primarily based on MoCA scores rather than a full neuropsychological assessment. We are fully aware that the clinical threshold of 26/30 is debated in the scientific community (Carson et al., 2018); however, this helped us demonstrate good sensitivity compared with CPP.

Overall, the present findings suggest that EasyCog is a promising digital tool enabling individuals and patients to quickly self-assess their cognitive functioning. Its self-administration facilitates user engagement and ownership of the tool while providing a detailed cognitive profile supported by robust psychometric properties. Furthermore, its capacity to detect subtle and potentially disease-specific cognitive alterations could make EasyCog a valuable asset for both clinical practice and research in chronic pain populations in the near future.

## Supporting information

screenshots of the digital tool

## Data Availability

Data available upon request and stored in the university of Strasbourg datacenter as recommended by our legal authorities.

## Conflicts of interest

XF and GJ are the cofounders of Traboule Labs, the company developing EasyCOG.

## Acknowledgments

We thank the following people for their help in the development process: Meggane Melchior, Iris Trinkler, Julia Devanne, and Olivier Despres.

## Funding

This study was financed by recurring funds from the CNRS and the University of Strasbourg and by grants from the French National Research Agency (ANR) through the Programme d’investissements d’avenir (contract ANR-17-EURE-022, EURIDOL, Graduate School of Pain), from Région Grand Est (ClueDOL, Fonds de coopération régionale et de recherche). PP is a senior fellow of the Institut Universitaire de France. SL and LH received a PhD scholarship from EURIDOL graduate school of pain. We thank ITI-neurostra for their complementary financial support to LH (grant ANR-20-SFRI-0012).

## Notes

### Author Declarations

The study was approved by the University of Strasbourg ethics committee (agreement number 2022-44) and was conducted in accordance with the Declaration of Helsinki.

