## Supplementary material for "EasyCog: a fast and effective numerical tool for the self-assessment of cognitive functions": screenshots of the digital tool

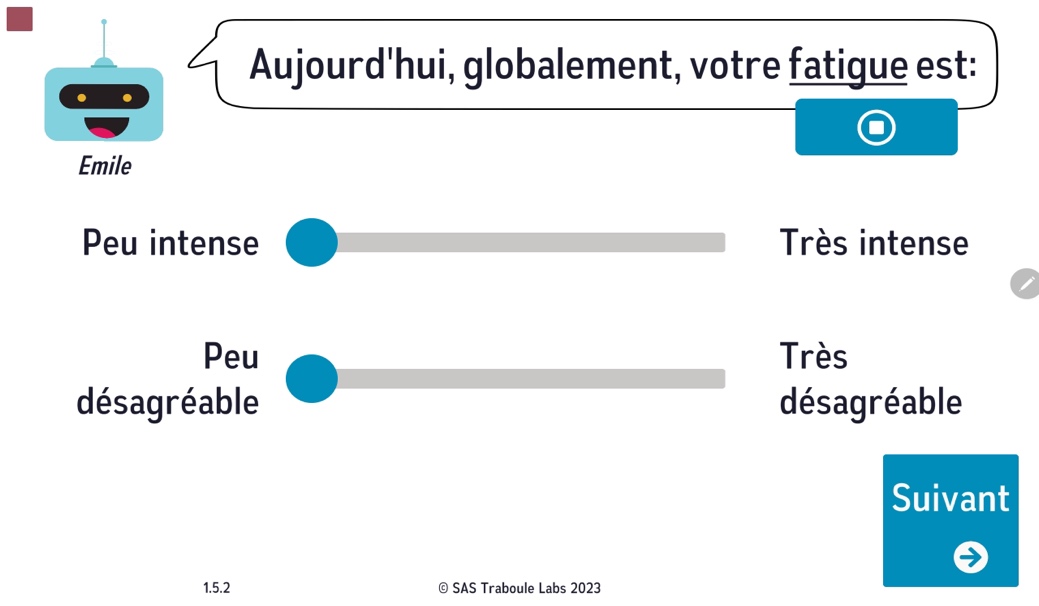

**Screenshot of the visual analogic scale** **(VAS)** for fatigue intensity and unpleasantness.

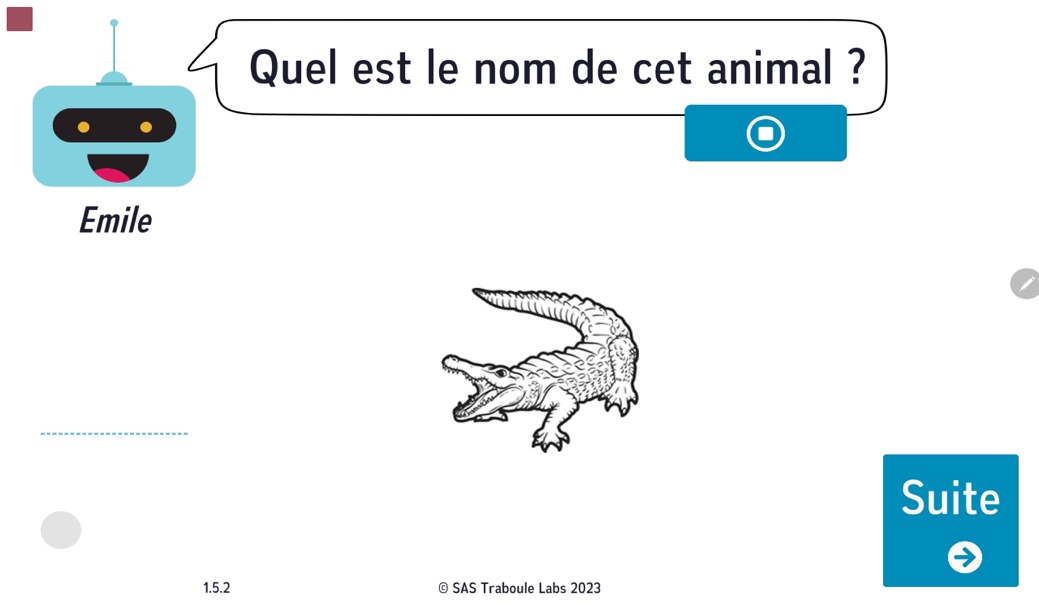

Emile chatbot

Voice recognition monitoring

« Next » button

**Screenshot of one of the naming test questions.**

Here the participant is asked to name verbally the drawing of an animal.

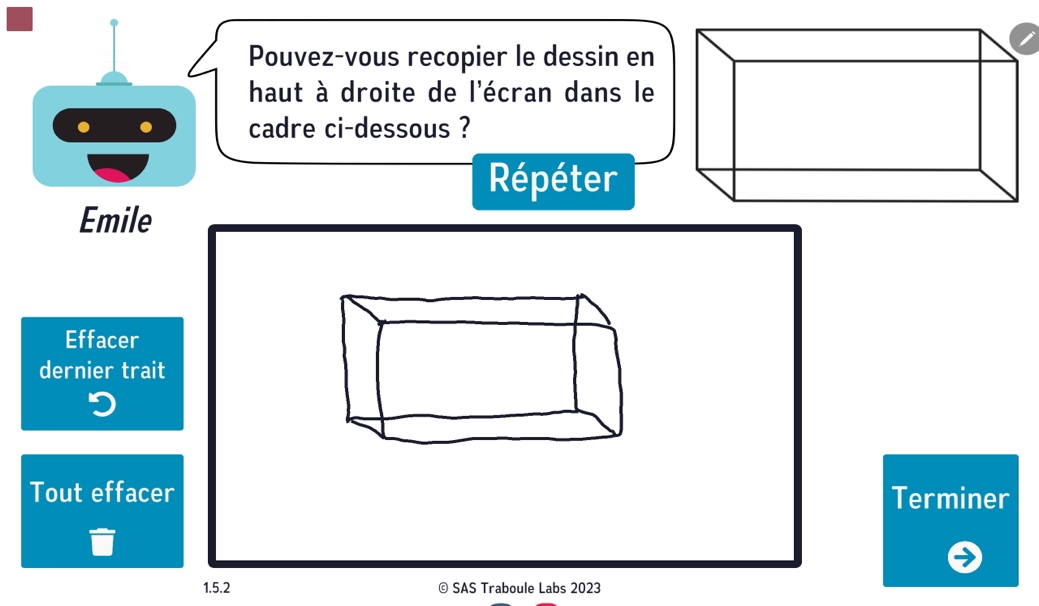

« Repeat » button

« Undo last stroke » button

« Clear all » button

« Finish » button

**Screenshot of the parallelepiped copy**. Here the participant is asked to reproduce the drawing of a parallelepiped as accurately as possible.
